# Hypothalamic structure and function in Alzheimer’s disease and Lewy-body dementia: a systematic review and meta-analysis

**DOI:** 10.1101/2024.09.03.24312999

**Authors:** Axel AS Laurell, Sita N Shah, Masoud Rahmati, John T O’Brien, Benjamin R Underwood

## Abstract

**Background:** Changes to sleep, weight, and endocrine function are common in Alzheimer’s disease (AD) and Lewy-body dementia (LBD). The cause of these is not known, but they may be related to hypothalamic neurodegeneration.

**Methods:** We performed a systematic search of MEDLINE and EMBASE of studies using structural magnetic resonance imaging to examine hypothalamic volume in people with AD or LBD. The Newcastle-Ottawa scale was used to assess the risk of bias. A random-effects meta-analysis was conducted using the standardised mean difference (SMD) in hypothalamic volume as the effect measure, and a narrative synthesis was used to examine the relationship between hypothalamic volume and sleep, eating, and endocrine function.

**Results:** We identified 6542 articles which resulted in 12 included studies, most which had a low to moderate risk of bias. The meta-analysis included 454 people with mild-moderate AD (Mini-Mental State (MMSE) range: 19.2-26.1) and 715 controls. We found that people with AD had a significantly smaller hypothalamus (pooled SMD: -0.49, t=-3.47, p=0.018, 95%CI: -0.86 to - 0.13). There was significant heterogeneity of a moderate degree (Tau^2^=0.0665, 95%CI:0.005-0.8090; I^2^=67%, 95%CI:21.5%-86.1%; Q=15.16, p<0.01), but no evidence of publication bias. Only one study examined people with LBD, finding qualitative evidence of lower hypothalamic volume compared to controls. Hypothalamic volume loss in AD was more marked in men and may be associated with plasma levels of sex hormones and decreased bone mineral density.

**Conclusion:** Reduced hypothalamic volume is seen early in AD and LBD and this may influence endocrine function. A better understanding of hypothalamic degeneration in dementia may help elucidate how pathology relates to symptoms in AD and LBD and reveal new targets for intervention.

## 1.0 Introduction

Alzheimer’s disease (AD) and Lewy-body dementia (LBD), consisting of Dementia with Lewy Bodies (DLB) and Parkinson’s disease dementia (PDD), are two of the most common causes of dementia (Goodman et al., 2017). In addition to the well-described cognitive impairment associated with these illnesses, many people with AD and LBD develop other symptoms including sleep disturbances (Koren et al., 2023), loss of appetite (Soysal et al., 2021), endocrine (Popp et al., 2015) and metabolic dysfunction (Xie et al., 2023). The causes of these are poorly understood, but may be related to neurodegenerative changes in specific brain regions that are involved in the control of these physiological processes. The hypothalamus is a small grey matter structure which consists of more than 12 different nuclei that are responsible for regulating a range of homeostatic, metabolic and autonomic functions, including appetite and the 24-hour circadian rhythm (Ishii and Iadecola, 2015). The hypothalamus is also the main regulator of many of the body’s endocrine processes via pathways such as the hypothalamic-pituitary-adrenal (HPA) axis, the hypothalamic-pituitary-gonadal (HPG) axis, the hypothalamic-pituitary-thyroid axis and the hypothalamic-pituitary-somatotropic axis. Changes in plasma levels of glucocorticoids (Huang et al., 2009), sex hormones (Livingston et al., 2024), thyroid hormones (Tang et al., 2021), and somatotropic hormones (Westwood et al., 2014) are associated with increased risk of developing dementia and metabolic conditions such as osteoporosis, indicating that hypothalamic signalling pathways may be involved (Niwczyk et al., 2023).

Although, neuropathological studies have established that the hypothalamus is affected by LBD (Braak et al., 2003) and AD pathology (Braak and Braak, 1991), it is not clear if this is an early or late feature of these conditions or whether there is a functional impact of neurodegeneration in this structure. These are important questions as it is possible that hypothalamic neuronal loss could drive global pathology, for example through changes in sleep, and any impact on the endocrine system could be related to AD symptomatology. The hypothalamus is a promising drug target due to its diverse and well understood receptor profile so understanding its role in disease might quickly translate into treatment (Romanov et al. 2017).

One challenge of studying the hypothalamus *in vivo* using magnetic resonance imaging (MRI) is its small size and poor tissue contrast, and traditional methods of manual segmentation are time-consuming and requires expert knowledge of neuroanatomy. Recently, several automatic segmentation pipelines have been developed which allows the hypothalamus to be segmented into subregions (Billot et al., 2020; Chang et al., 2022). This allows the hypothalamic volume to be evaluated in larger cohort studies to assess whether the hypothalamus is affected early in dementia. It also makes it possible to test the hypothesis that volumetric changes in specific hypothalamic subunits are associated with specific signs and symptoms. The primary outcome of this meta-analysis was to assess whether there was evidence for any structural changes in the hypothalamus in people with AD or LBD using *in vivo* structural MRI. The secondary objective was to assess whether there was an association between the structure of the hypothalamus and dementia symptoms including changes to weight, sleep and endocrine function.

## 2.0 Methods

The protocol for the review was preregistered on PROSPERO (https://www.crd.york.ac.uk/prospero/, registration number: CRD42024547743) and the manuscript was produced using the PRISMA guidelines (Page et al., 2021).

### 2.1 Search strategy

We performed a systematic search of MEDLINE and EMBASE from inception to 15^th^ May 2024 using the Ovid platform, to identify studies of people with AD or LBD which examined the structure of the hypothalamus. The search string used was (hypothal* AND (MRI or magnetic resonance imaging) AND (Alzheimer* OR DLB OR Lewy OR LBD OR Parkinson* disease dementia OR PDD)). Filters were used to limit the search to human studies in English, using people aged 18 or over, published in “journal” or “report”, and to remove conference abstracts and duplicates.

### 2.2 Eligibility criteria

We only included studies using structural MRI imaging *in vivo* in humans, assessing the volume of the hypothalamus, and excluded neuropathological studies and studies of animals. Case studies and series, reviews, meta-analyses, conference abstracts and non-peer reviewed material were also excluded.

### 2.3 Data extraction and quality assessment

Two authors (AASL and SNS) worked independently to screen the titles, abstracts and full-texts to identify articles for inclusion using the online platform Rayyan (rayyan.ai). The reference lists of all included studies were reviewed to identify any further studies meeting the inclusion criteria. Any disagreements were resolved by discussion with a third reviewer (BRU). The Newcastle-Ottawa scale for case-control studies was used to assess the quality and risk of bias of all included studies (Wells et al., 2021). This tool assesses non-randomised studies using three domains (selection, comparability and exposure), to allow studies to be stratified as being of “low” (8-9 points), “moderate” (6-7 points) or “high” (≤ 5 points) risk of bias. Data was extracted from the included studies using pre-defined tables. For the meta-analysis, this included the mean hypothalamic volume, standard deviation and sample size of the AD and control groups. For the narrative synthesis, this included any qualitative evidence of group difference in hypothalamic volume (from whole brain voxel-based morphometry (VBM) studies), and associations between hypothalamic volume, weight or appetite, sleep and endocrine function. Two studies met the inclusion criteria but did not report sufficient quantitative data to be included in the meta-analysis. After contacting the authors of both studies, one was able to provide the necessary additional data (Ofori et al., 2024), whereas the other study was included in the narrative synthesis but not the meta-analysis (Billot et al., 2020).

### 2.4 Statistical analysis

A random-effects meta-analysis was conducted using the meta package in R studio (version 2021.09.0) to determine whether there was a difference in hypothalamic volume between dementia and controls. The standardised mean difference (SMD, Cohen’s d) between AD and controls was calculated to homogenise the data, since studies reported volumetric changes in the hypothalamus in different units (mm^3^ or % of intracranial volume). Hedges’ g was then calculated and used as the effect size in the meta-analysis. This applies a small-sample correction factor to the Cohen’s d, since the effect size can be overestimated in studies with small sample sizes. A Hedges’ g of 0.2 indicates a small effect, while 0.5 and 0.8 indicates a medium and large effect respectively. The percentage difference in volume for each study was also calculated and averaged across the number of included studies since this may be more easily interpretable. Heterogeneity (Tau^2^) was estimated using the restricted maximum likelihood procedure, and Knapp-Hartung adjustment was applied to control for the uncertainty of the between-study heterogeneity. The Q statistic was calculated to assess whether the level of between-study heterogeneity was significant and the I^2^ statistic was used to quantify this (25%= low, 50%=moderate, 75%=substantial) (Higgins and Thompson, 2002). Publication bias was assessed by Egger’s regression test, and by inspection of the funnel plot. A narrative synthesis strategy was applied to assess whether there was a relationship between structural changes and non-cognitive symptoms of dementia.

## 3.0 Results

The article selection process is displayed in Figure 1. The search identified the titles and abstracts of 6542 articles which were screened to identify 63 articles for full-text review after being marked for further consideration by at least one author. A list of the articles excluded after full-text review is available in supplementary Table S1. Two articles were identified from screening the reference lists of included studies and relevant review articles. This resulted in the inclusion of 6 articles which provided quantitative measurements of hypothalamic volume in AD that could be extracted for the meta-analysis (Grundman et al., 1996; Callen et al., 2001; Loskutova et al., 2010; Ahmed et al., 2015; Chang et al., 2023; Ofori et al., 2024). A further 6 studies reported qualitative changes in hypothalamic volume as part of whole brain VBM, and/or associations between hypothalamic volume and clinical symptoms, and these were included in the secondary narrative synthesis (Baron et al., 2001; Callen et al., 2004; Whitwell et al., 2007; Hall et al., 2008; Kim et al., 2016; Billot et al., 2020). There was only one study examining people with DLB using VBM, and no studies which included people with PDD. The included studies had mostly (83%) a low, or moderate risk of bias, with only two studies rated as high risk of bias. The extracted data is displayed in Table 1.

**Figure 1.**
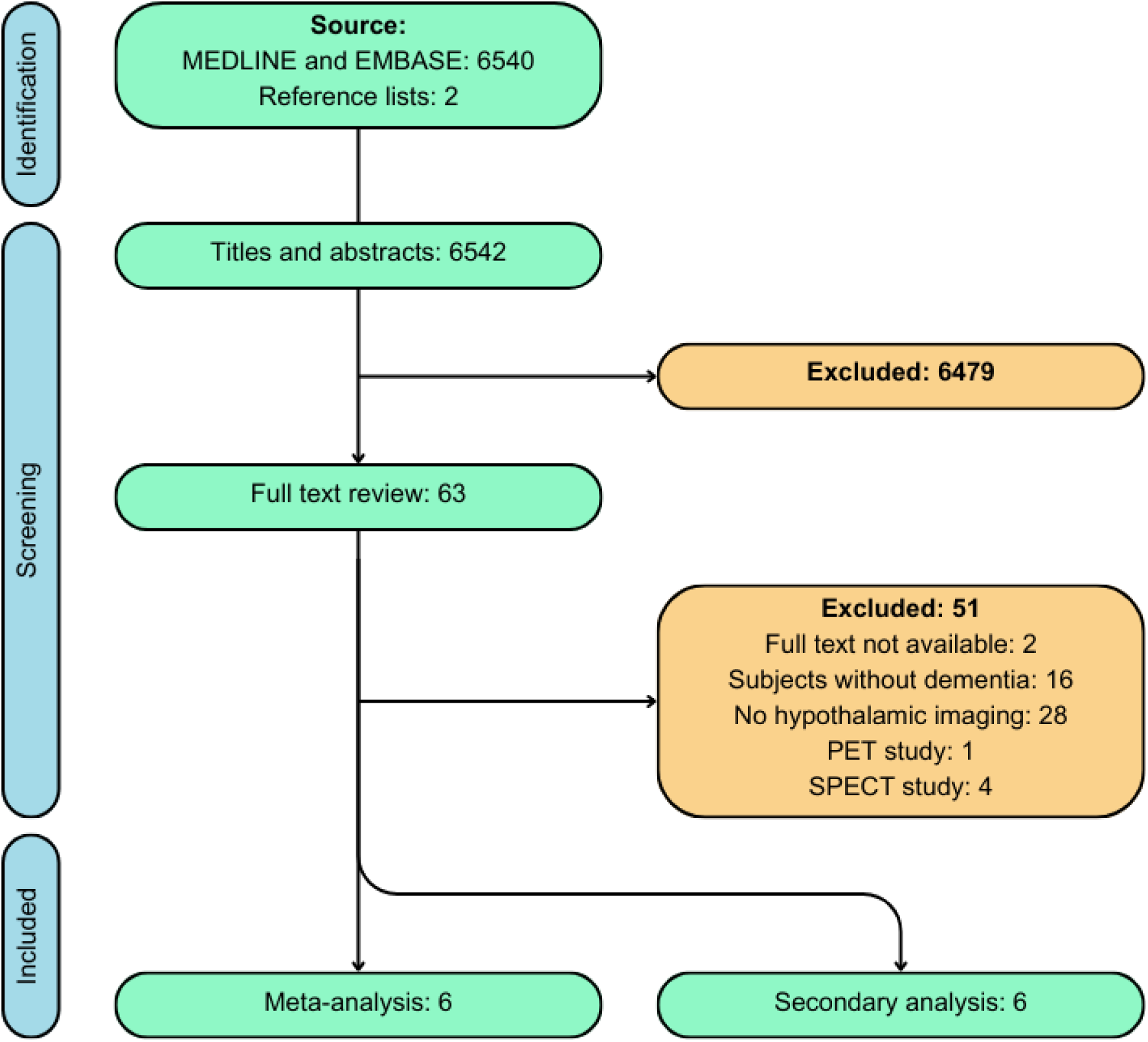
Flowchart showing the selection of articles for the meta-analysis and secondary narrative synthesis. PET = positron emission tomography, SPECT = single-photon emission computed tomography.

**Table 1.** Summary of studies included in the data synthesis. ACE-R = Addenbrooke’s cognitive examination revised, AD = Alzheimer’s disease, Ant-Inf = anterior-inferior subunit, Ant-Sup = anterior-superior subunit, APEHQ = Appetite and eating habits questionnaire, BMI = body mass index, CBI = Cambridge behavioural inventory, DLB = dementia with Lewy bodies, FSH = follicle-stimulating hormone, Inf-Tub = inferior-tubular subunit, LH = lutenising hormone, M = mini-mental state examination, 3MS = modified mini-mental state examination, MPRAGE = magnetization prepared rapid gradient echo, NS = not significant, OSHy-X = open source hypothalamic-fornix atlas and tool, Post – posterior subunit, RBDSQ-K = Rapid eye movement behaviour disorder screening questionnaire Korean, ROIs = region of interests, SD = standard deviation, SPGR = spoiled gradient echo, Sup-Tub = superior-tubular subunit, T = Tesla, TIV = total intracranial volume, VBM = voxel based morphometry, %W = % women.

| Study<br>Author<br>(year) | Demographics<br>Group (n)<br>Age, %W<br>Cognition: M, 3MS, or ACE-R | MRI characteristics<br>Field strength<br>Sequence<br>Voxel or slice size<br>Analysis method | Risk of<br>bias | Hypothalamic volume<br>Mean (SD) | Mean difference AD<br>compared to controls<br>(%) | Clinical correlations |
| --- | --- | --- | --- | --- | --- | --- |
| Grundman<br>(1996) | AD (58)<br>71.8y, 50%W, M:19.2<br>Controls (16)<br>72.5y, 69%W, M:28.9 | 1.5T<br>Multiple-echo sequence.<br>Slices: 5mm.<br>Manual ROIs. | Low | AD<br>0.12% (0.05%) of TIV<br>Controls<br>0.17% (0.04%) of TIV | -29.41% | BMI: r=0.14 (NS) |
| Baron<br>(2001) | AD (19)<br>73y, 58%W, M: 22.4<br>Controls (16)<br>66y, 56%W, M not reported | 1.5T<br>T1 (SPGR) sequence<br>Voxels: 2mm<sup>3</sup><br>VBM | Moderate | Gray matter loss in anterior<br>hypothalamus in AD<br>compared to controls. | Not measured. | Not measured. |
| Callen<br>(2001) | AD (40)<br>69.1y, 50%W, M:19.5<br>Controls (40)<br>70.4y, 50%W, M:28.5 | 1.5T<br>T1 sequence.<br>Voxels: 0.634 mm<sup>3</sup>.<br>Manual ROIs. | Low | AD<br>266.5 (54.3) mm<sup>3</sup><br>Controls<br>298.6 (43.2) mm<sup>3</sup> | -10.75% | Not measured |
| Callen<br>(2004) | Same as Callen (2001). | Same as Callen (2001). | Low | Same as Callen (2001). | Same as Callen (2001). | Sex (men<women) |
| Whitwell<br>(2007) | AD (73)<br>76y, 24%W, M:21<br>DLB (73)<br>73y, 24%W, M:22<br>Controls (73)<br>74y, 24%W, M:29 | 1.5T<br>T1 (SPGR) sequence<br>Slices: 1.6mm<br>VBM | Low | Grey matter loss in the<br>hypothalamus in DLB<br>compared to controls. | Not measured. | Not measured. |
| Hall<br>(2008) | AD (26)<br>83.2y, 61%W, 3MS:76.7<br>Controls (127)<br>77.3y, 54%W, 3MS:96.9 | 1.5T<br>T1 (SPGR) sequence<br>Voxels: 1 mm<sup>3</sup><br>VBM | Moderate | Grey matter loss in the<br>hypothalamus in AD<br>compared to controls, and<br>in subjects who later<br>developed AD. | Not measured. | Not measured. |

Table 1 continued.
| Study<br>Author<br>(year) | Demographics<br>Group (n)<br>Age, %W<br>Cognition: M or ACE-R | MRI characteristics<br>Field strength<br>Sequence<br>Voxel or slice size<br>Analysis method | Risk of<br>bias | Hypothalamic volume<br>Mean (SD) | Mean difference AD<br>compared to controls<br>(%) | Clinical correlations |
| --- | --- | --- | --- | --- | --- | --- |
| Loskutova<br>(2010) | <u>AD (63)</u><br>74.4y, 62%W, M:26.1<br><u>Controls (55)</u><br>73.2y, 58%W, M:29.5 | 3T<br>T1 (MPRAGE) sequence<br>Voxels: 1 mm<sup>3</sup>.<br>VBM with small volume correction. | Moderate | <u>AD</u><br>570 (100) mm<sup>3</sup><br><u>Controls</u><br>650 (120) mm<sup>3</sup> | -12.31% | Bone mineral density: r = 0.34,<br>(p = 0.007) |
| Ahmed<br>(2015) | <u>AD (30)</u><br>68y, 27%W, ACE-R: 68<br><u>Controls (23)</u><br>72y, 52%W, ACE-R: 94 | Field strength not stated.<br>T1 sequence<br>Voxels: 1 mm<sup>3</sup>.<br>Manual ROIs. | Moderate | <u>AD</u><br>0.138% (0.02%) of TIV.<br><u>Controls</u><br>0.137% (0.013%) of TIV | +0.73% | CBI (NS), APEHQ (NS)<br>BMI (NS)<br>Leptin (NS)<br>Cholecystokinin (NS)<br>Peptide YY (NS)<br>Oxytocin (NS)<br>Ghrelin (NS)<br>Agouti-related peptide (NS) |
| Kim<br>(2016) | <u>AD (23)</u><br>71.6y, 61%W, M:20,1<br><u>Controls (36)</u><br>72.9y, 47%W, M:29.3 | 3T<br>T1 (SPGR) sequence.<br>Slices: 1mm.<br>VBM | Low | Not reported. | Not reported. | RBDSQ-K score (trend only). |
| Billot<br>(2020) | <u>AD (134)</u><br>76.0y, 46%W<br><u>Controls (183)</u><br>72.9y, 49%W<br>M not reported. | T1<br>Sequence not reported.<br>Voxels: 1mm<sup>3</sup>.<br>Automated segmentation using Freesurfer. | High | Not reported. | <u>Cohen's d</u><br>Hypothalamus (0.96)<br>Ant-Sup (1.16)<br>Ant-Inf (0.99)<br>Sup-Tub (0.60)<br>Inf-Tub (0.34)<br>Post (0.94) | Not reported. |
| Chang<br>(2022) | <u>AD (167)</u><br>68y, 50%W<br><u>Controls (527)</u><br>68y, 58%W<br>M not reported | 3T or 7T<br>T1 (MPRAGE or accelerated inversion recovery<br>SPGR) sequence<br>Voxels: 1mm<sup>3</sup> or 0.9 mm<sup>3</sup>.<br>Automated segmentation using OSHy-X. | High | <u>AD</u><br>746.62 mm<sup>3</sup> (121.29)<br><u>Controls</u><br>771.2 mm<sup>3</sup> (97.88) | -3.19% | BMI (NS)<br>Sex (NS) |
| Ofori<br>(2024) | <u>AD (96)</u><br>75.1y, 45%W, M:21.3<br><u>Controls (54)</u><br>75.3y, 48%W, M:29.1 | 1.5T<br>T1 (MPRAGE) sequence.<br>Automatic segmentation using Freesurfer. | Moderate | <u>AD</u><br>0.034% (0.0039%)<br><u>Controls</u><br>0.036% (0.0037%) | -5.56% | Sex (men<women)<br><u>In women only:</u><br>FSH and Inf-Tub. (R<sup>2</sup>=0.11)<br>LH and Inf-Tub. (R<sup>2</sup>=0.11)<br>Testosterone (NS)<br>Progesterone (NS) |

One study which initially appeared to meet the inclusion criteria, demonstrated that the volume of the mamillary bodies, which is part of the posterior hypothalamus, was reduced in people with AD compared to controls (Copenhaver et al., 2006). This was not included in the data synthesis since it did not provide a volumetric measure for the full hypothalamus, although it is consistent with the findings of the included studies.

### 3.1 Hypothalamic volume in AD and DLB versus controls

The results of the random-effects meta-analysis is displayed in Figure 2, which includes data from a total of 454 people with mild-moderate AD (MMSE range: 19.2-26.1) and 715 healthy controls. The pooled standardised mean difference was -0.49 (t=-3.47, p=0.018, 95%CI: - 0.86 to -0.13), indicating that people with AD had a significantly smaller hypothalamus, a difference which can be considered to be of a small-to-medium effect size. The between-study heterogeneity variance was estimated at Tau^2^ = 0.0665 (95%CI: 0.005-0.8090), I^2^ = 67% (95%CI: 21.5%-86.1%) and Q = 15.16 (p<0.01). This indicates that there was significant heterogeneity to a moderate degree. On average, the volume of the hypothalamus was reduced by 10.1% (range: -29.4% to +0.73%) in people with AD compared to controls. It is important to highlight that no study normalised their findings to the total grey matter volume, making it difficult to know whether the difference in hypothalamic volume is over and above the general grey matter atrophy which is seen in AD. The funnel plot (Figure 3) was largely symmetrical and revealed no signs of publication bias. Egger’s regression test was not significant (intercept -2.16, p=0.23).

**Figure 2.**
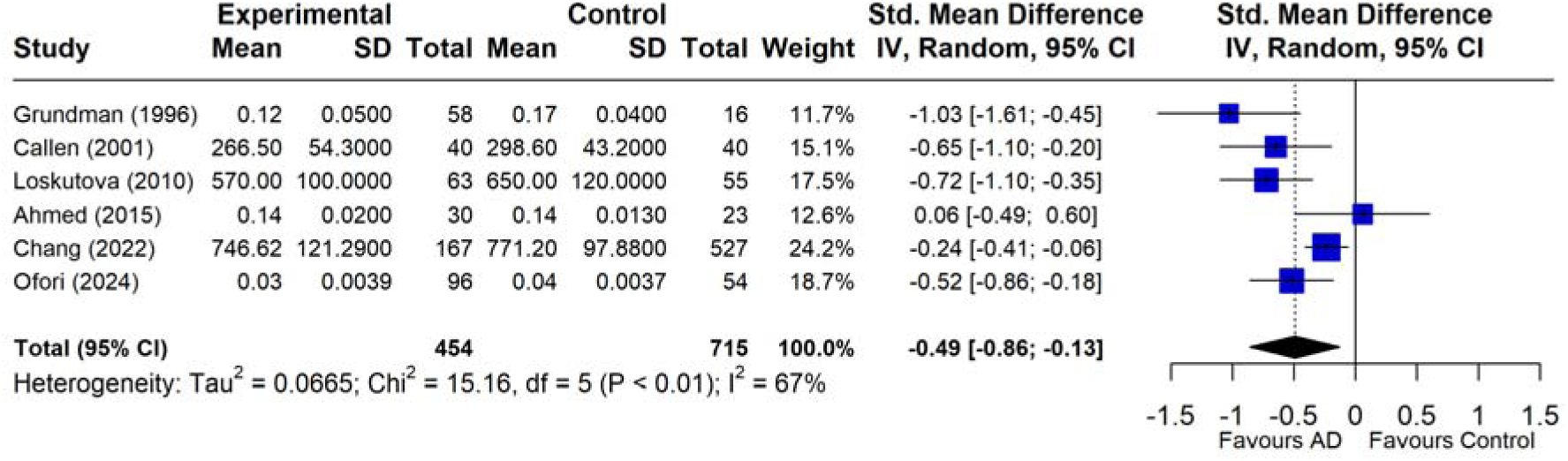
Forest plot of the pooled standardised mean difference (Hedges’ g) showing that subjects with Alzheimer’s disease (AD) had a significantly smaller hypothalamus compared to healthy controls.

**Figure 3.**
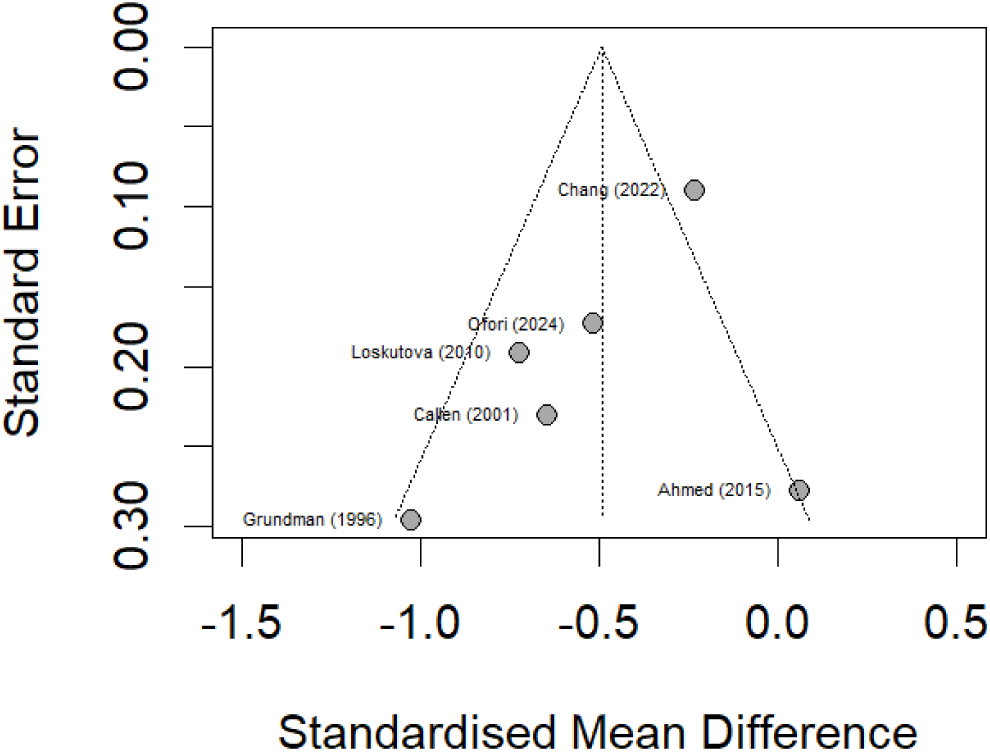
Funnel plot of included studies demonstrating no suggestion of publication bias.

Four studies used whole-brain VBM to provide a qualitative measure of differences in voxels between people with dementia and controls. Three of these reported grey matter loss in the hypothalamus in people with AD (Baron et al., 2001; Whitwell et al., 2007; Hall et al., 2008), while one did not report any differences (Kim et al., 2016). The only study using VBM in people with DLB found grey matter loss in the hypothalamus in DLB compared to controls (Whitwell et al., 2007). Billot et al. used an automatic segmentation pipeline and found reduced volumes of all hypothalamic subunits in a large group of people with MCI and AD compared to healthy controls, but did not provide sufficient volumetric data for inclusion in the meta-analysis (Billot et al., 2020). The group differences were particularly pronounced in the anterior-superior, anterior-inferior and the posterior hypothalamic subunits (Cohen’s d = 1.16, 0.99, 0.94 respectively), with a relative sparing of the inferior-tubular subunit (Cohen’s d = 0.34). Although, these studies are consistent with the results of the meta-analysis, there may be other VBM studies comparing AD to healthy controls which did not specifically mention the hypothalamus (due to a lack of an effect), and which were therefore not captured by our search string.

### 3.2 Hypothalamic volume in AD versus other neurodegenerative conditions

There was no difference in hypothalamic volume in people with AD compared to DLB (Whitwell et al., 2007), amyotrophic lateral sclerosis (ALS) (Chang et al., 2023), or Parkinson’s disease (Kim et al., 2016). One study found that people with behavioural variant frontotemporal dementia (bvFTD) had a significantly smaller hypothalamus compared to AD and controls (Ahmed et al., 2015). However, in this study the hypothalamic volume for the AD group were not different from the healthy controls, making the group differences difficult to interpret.

### 3.3 Hypothalamus and weight

There was no association between hypothalamic volume and body mass index (BMI) (Grundman et al., 1996; Ahmed et al., 2015; Chang et al., 2023), or with eating habits as measured by the Cambridge Behavioural Inventory (CBI) or the Appetite and Eating Habits Questionnaire (APEHQ) (Ahmed et al., 2015). In all three studies the AD group had normal or slightly raised BMI (mean: 23.8, 26.4, 26.6).

### 3.4 Hypothalamus and sleep

Higher score on the rapid eye movement (REM) sleep behaviour disorder screening questionnaire, indicating symptoms of REM sleep behaviour disorder, was associated with grey matter volume in the hypothalamus, although the association disappeared when correcting for family-wise error (Kim et al., 2016).

### 3.5 Hypothalamus and endocrine function

Larger hypothalamic volume was associated with increased bone mineral density in AD (r=0.34, p=0.007) (Loskutova et al., 2010). There was no significant association between hypothalamic volume and plasma levels of appetite-regulating hormones, including leptin, cholecystokinin, neuropeptide Y, agouti-related peptide, and peptide YY (Ahmed et al., 2015). Two studies reported that hypothalamic volume was smaller in men than women with AD (Callen et al., 2004; Ofori et al., 2024), while one study with a high risk of bias found no effect of sex (Chang et al., 2023). This effect was seen despite the males in the AD group being at an earlier stage of illness than the females in one of the studies (Callen et al., 2004). Ofori et al. reported several associations between hypothalamic volume and plasma levels of follicle stimulating hormone (FSH), luteinizing hormone (LH), progesterone and testosterone, although these were not corrected for multiple comparisons (Ofori et al., 2024). Increased volume of the inferior-tuberal subunit was weakly associated with levels of FSH and LH in women with AD (R^2^=0.11 (FSH), 0.12 (LH)), while no associations were found in men with AD. In people with mild cognitive impairment (MCI), there were weak associations between FSH, LH, progesterone, and testosterone and the volume of all hypothalamic subunits apart from the anterior-inferior subunit (R^2^=0.05-0.08). In healthy controls there were associations between the inferior-tuberal subunit and progesterone (R^2^=0.18 (men) and R^2^=0.38 (women)), inferior-tuberal subunit and testosterone (R^2^=0.20 women only), superior-tuberal subunit and testosterone (R^2^=0.20 men only), and posterior hypothalamus and LH (R^2^=0.21 men and R^2^=0.21 women. This suggests that the association between hypothalamic volume and plasma sex hormones weakens with worsened cognitive status.

In summary, multiple studies support that people with AD have a significantly lower hypothalamic volume than controls. There has been one study in people with DLB with similar findings. Furthermore, sex may influence hypothalamic volume loss, and decreased hypothalamic volume may be associated with lower bone mineral density and plasma levels of sex hormones although this needs further confirmation. No significant associations were found with BMI or sleep.

## 4.0 Discussion

We found that people with AD have a smaller hypothalamus compared to healthy controls, as measured by *in vivo* volumetric MRI imaging. Most of the studies investigated people with mild-to-moderate AD, indicating that hypothalamic changes are present early in the clinical course. A previous narrative review has described the role of the hypothalamus in AD, although it only included evidence from 7 volumetric MRI studies (Ishii and Iadecola, 2015). Here, we use both a meta-analytical and narrative approach to synthesise the evidence from 12 studies on the structure and function of the hypothalamus in AD and DLB, including studies which uses recently developed automatic segmentation pipelines (Billot et al., 2020; Chang et al., 2023; Ofori et al., 2024). The findings are consistent with neuropathological studies which have found amyloid beta plaques and tau neurofibrillary tangles throughout the hypothalamus in people with AD, associated with a reduced number of neurons and morphological changes to organelles (Baloyannis et al., 2015; Braak and Braak, 1991; Lim et al., 2014). Similarly, Lewy-body pathology has also been found in the hypothalamus of people with PD, DLB and PDD (Benarroch et al., 2015; Braak et al., 2003; Kasanuki et al., 2014; Purba et al., 1994).

Our results are also consistent with studies using other MRI modalities, which have found reduced glucose metabolism in the hypothalamus in people with MCI and AD using positron emission tomography (Cross et al., 2013; Liguori et al., 2017; Nestor et al., 2003). One study using single-photon emission computed tomography (SPECT) with the tracer ^123^I-N-ω-fluoropropyl-2β-carbomethoxy-3β-(4-iodophenyl)nortropane (^123^I-FP-CIT) found that people with DLB had lower serotonergic and/or dopaminergic binding in the hypothalamus compared with healthy controls (Joling et al., 2019), although other SPECT studies in AD have failed to demonstrate reduced perfusion in the hypothalamus (Callen et al., 2002, 2004; Ismail et al., 2008; Lanctot et al., 2004, 2007). This discrepancy is likely a reflection of the resolution of a SPECT scan being too low to detect any changes in a small structure such as the hypothalamus without a specific tracer.

Studies in other neurodegenerative conditions using both manual and automatic segmentation protocols have shown that the hypothalamus is smaller in all subtypes of FTD (9-17% reduction) (Piguet et al., 2011; Ahmed et al., 2015; Bocchetta et al., 2015; Shapiro et al., 2022; Tse et al., 2023), ALS (22% reduction) (Gorges et al., 2017; Liu et al., 2022; Tse et al., 2023), in pre-symptomatic carriers of pathogenic FTD-ALS mutations (14% reduction) (Gorges et al., 2017), and in spinocerebellar ataxia type 3 (3.40% reduction) (Guo et al., 2022). A VBM study in people with Huntington’s disease, suggested that hypothalamic changes were present 9-15 years before the predicted time of clinical diagnosis (Soneson et al., 2010), although another study using manual segmentation failed to find any cross-sectional or longitudinal group differences (Gabery et al., 2015). Studies comparing different types of dementia do not suggest that the magnitude of volume loss is significantly different to that seen in AD and DLB. This suggests that the hypothalamus is vulnerable to neurodegeneration secondary to a range of different pathologies. It is less well understood whether the loss of volume in the hypothalamus contributes to the development of dementia, symptomatology or whether this might be amenable to therapeutic intervention.

### 4.1 Hypothalamus and weight

Physiologically, appetite is mediated by the hormones leptin and ghrelin, which is secreted peripherally by adipose tissue and the stomach respectively (Coll et al., 2007). They have opposing actions on the signalling between the arcuate and the paraventricular nuclei (PVN) of the hypothalamus, to promote either satiety or increased appetite. Several studies have assessed this pathway in people with bvFTD, which is commonly associated with binge-eating, a preference for sucrose and weight gain (Bang et al., 2015). People with bvFTD overeat despite having a reduced plasma levels of ghrelin and raised leptin, indicating that there is a central resistance to the effect of these hormones (Woolley et al., 2014). While people with bvFTD had reduced volume of most hypothalamic subunits, there was a relative sparing of the inferior-tubular subunit, which contains the arcuate nucleus (Shapiro et al., 2022; Tse et al., 2023). Furthermore, BMI and eating behaviour, as measured by CBI-R, is negatively associated with the volume of the anterior-superior, anterior-inferior and posterior subunits in bvFTD and ALS but not with the inferior-tubular subunit (Gorges et al., 2017; Liu et al., 2022; Shapiro et al., 2022; Tse et al., 2023). These findings suggests that the dysregulated appetite in bvFTD is not due to changes in the signalling to the arcuate nucleus but may instead be due to changes in higher order nuclei, such as the PVN, or in other areas of the brain. Interestingly, the inferior-tubular subunit was relatively preserved in AD on MRI imaging (Billot et al., 2020), while neuropathological studies have found that in this nucleus is either unaffected by pathology (Purba et al., 1994; Saper and German, 1987), or only affected by pathology in men (Schultz et al., 1996, 1999). This suggests that the arcuate nucleus may be more resistant to neurodegeneration than other hypothalamic nuclei. Weight loss is common in both AD and LBD, and this is associated with an increased rate of mortality and emergency hospitalisation (Soysal et al., 2021). Although there was no association between appetite or BMI and hypothalamic volume in AD (Grundman et al., 1996; Ahmed et al., 2015; Chang et al., 2023), this may be due to the normal or slightly raised BMI of people with AD in these studies. Longitudinal studies are needed to assess whether weight loss is related to hypothalamic volume in AD or DLB, and whether the subunit-specific associations are similar to those observed in bvFTD.

### 4.2 Hypothalamus and sleep

Sleep disturbance is very common in people living with dementia, with an estimated prevalence of 24% in Alzheimer’s disease and 49% in Lewy body dementia (Koren et al., 2023). This is a significant issue both for people with dementia and their caregivers where sleep disturbance is associated with carer stress and need for institutional care. Although the underlying causes are likely to be multifactorial, it is possible that it involves neurodegeneration of the hypothalamus, since this structure is essential for the regulation of sleep and wakefulness. Wakefulness is promoted by the histaminergic tuberomammillary nucleus (TMN) in the posterior hypothalamus, while sleep is promoted by the GABA and galanin producing neurons of the ventrolateral preoptic nucleus (VLPO) in the anterior hypothalamus (Saper et al., 2005). The lateral hypothalamus modifies this system through orexin, which promotes wakefulness, and melanin-concentrating hormone, which promotes sleep. The 24-hour circadian rhythm is maintained by the suprachiasmatic nucleus (SCN) in the anterior-inferior hypothalamus, which influences the VLPO and lateral hypothalamus via its connections to the dorsomedial nucleus. Disruptions to this complex system can cause sleep-disorders, such in the case of narcolepsy which is characterized by a selective loss of orexin-producing neurons (Saper et al., 2005). Neuropathological studies have shown that people with DLB and AD had fewer neurons in the VLPO, SCN and TMN compared to controls, with associations between the number of neurons and sleep fragmentation, circadian rhythm and core body temperature (Harper et al., 2008; Fronczek et al., 2012; Shan et al., 2012; Kasanuki et al., 2014; Lim et al., 2014).

In contrast, *in vivo* studies have found that people with MCI and AD have increased levels of CSF orexin compared with controls, with an association between orexin levels and disrupted sleep metrics (Liguori et al., 2014, 2016, 2017; Gabelle et al., 2017). In DLB and PDD, studies have failed to demonstrate any alterations in CSF orexin or any relationship between orexin and changes in sleep (Yasui et al., 2006; Compta et al., 2009). Furthermore, treatment with suvorexant, a dual orexin receptor antagonist, have received Food and Drug Administration (FDA) approval for the treatment of insomnia in Alzheimer’s disease after it was shown to increased total sleep time and reduce waking after sleep onset compared to placebo (Herring et al., 2020). Suvorexant also acutely reduce the concentration of CSF amyloid beta and ptau181/tau181 ratio in healthy volunteers (Lucey et al., 2023), and a longitudinal trial in people at risk of AD is ongoing (https://clinicaltrials.gov/study/NCT04629547). Only two small MRI studies have examined association between hypothalamus and sleep in AD, finding a significant association between reduced hypothalamic glucose uptake and increased sleep disturbances (Liguori et al., 2017), and a trend towards higher REM sleep behaviour disorder score and grey matter signal on VBM (Kim et al., 2016). Further studies will need to clarify how a reduction in hypothalamic volume relates to an increase in CSF orexin in AD.

### 4.3 Hypothalamus and endocrine function

In addition to maintaining weight, appetite and sleep, the hypothalamus is an important regulator of several endocrine glands. The HPA axis starts with secretion of corticotrophin-releasing hormone (CRH) from the PVN, which in turn stimulates secretion of adrenocorticotropic hormone (ACTH) from the pituitary gland (Ahmad et al., 2019). This stimulates the secretion of glucocorticoids, such as cortisol, from the adrenal glands which acts through a negative feedback loop on the hypothalamus and pituitary gland to limit its own secretion. People with MCI and AD have higher levels of cortisol in plasma and CSF (Csernansky et al., 2006; Popp et al., 2015), an abnormal dexamethasone suppression test (O’Brien et al., 1996; Elgh et al., 2006), and increased CRH production in the PVN compared to healthy controls (Raadsheer et al., 1995). Furthermore, these abnormalities were associated with higher cerebral amyloid burden (Toledo et al., 2012), increased hippocampal atrophy (O’Brien et al., 1996; Elgh et al., 2006; Huang et al., 2009), and more rapid decline on cognitive tests (Huang et al., 2009; Popp et al., 2015), indicating that dysfunction of the HPA axis may contribute to the progression of the disease.

Similarly, the HPG axis consists of gonadotrophin-releasing hormone (GnRH) which is secreted in a pulsatile manner from the arcuate nucleus of the hypothalamus to stimulate the secretion of follicle-stimulating hormone (FSH) and luteinising hormone (LH) from the anterior pituitary gland (Desai et al., 2021). FSH and LH in turn stimulate the release of sex hormones (testosterone, oestrogens and progesterone) from the gonads which acts through a negative feedback loop on the hypothalamus and pituitary to regulate the HPG axis. The decline in plasma levels of neuroprotective sex hormones after menopause, may explain why AD is more prevalent in women than men (Livingston et al., 2024). However, a meta-analysis of randomised clinical trials found that hormone replacement therapy with oestrogen and progesterone after menopause was associated with an increased risk of dementia, although the timing of initiation, type of formulation and duration of treatment may modify this risk (Zhou et al., 2021). It is also possible that an increase in GnRH, FSH and LH which also occurs because of menopause may contribute to the increased risk of AD in women. Increased levels of FSH and LH, were associated with higher amyloid load, reduced grey matter volume and hippocampal loss independent of age and menopause status (Nerattini et al., 2023).

A similar pattern is seen in men with AD, who have lower levels of testosterone and higher levels of FSH and LH compared to controls (Bowen et al., 2000; Butchart et al., 2013). One ecological study suggests that men who have had treatment with an GnRH agonists for prostate cancer, had a lower risk of developing AD compared to people who underwent other procedures (Smith et al., 2018). GnRH agonists work by continuously stimulating the GnRH receptor in the pituitary which initially causes a spike in FSH, LH and testosterone production, followed by downregulation of GnRH receptors in the pituitary gland (Desai et al., 2021). The result is a reduction of FSH, LH and testosterone. The only study using hypothalamic segmentation to investigate the HPG axis in AD, found that the volume of the inferior tubular subunit, which contains the arcuate nucleus, was associated with plasma levels of FSH and LH in women, but not men, with AD (Ofori et al., 2024). Furthermore, in healthy controls and people with MCI, there was a more widespread associations between hypothalamic volume and plasma levels of FSH, LH, progesterone and testosterone. The hypothalamus is one of the most highly sexually dimorphic areas of the brain, with healthy men having a larger hypothalamus than women (Goldstein et al., 2001). Conversely, two studies in the current review found that men with AD have a smaller hypothalamus than women (Callen et al., 2004; Ofori et al., 2024), a finding which is consistent with the neuropathological studies of the arcuate nucleus which is almost exclusively affected by AD pathology in men but not women (Schultz et al., 1996, 1999). Overall, these studies provide further indication that hypothalamic pathology may be relevant to explain the sex discrepancy seen in AD.

Hypothalamic pathology in AD may also have an impact on other hormones which are secreted by the hypothalamus, including growth-hormone releasing hormone (GHRH) and thyrotropin releasing hormone (TRH). In people with MCI, treatment with GHRH analogues improves executive function and levels of insulin-like-growth factor 1 (Baker et al., 2012), and lower plasma levels of this hormone are associated with an increased risk of developing AD (Westwood et al., 2014). Furthermore, a meta-analysis found that a both an increased levels of thyroid stimulating hormone (TSH) (indicating primary hypothyroidism) and reduced TSH levels (indicating primary hyperthyroidism) were associated with an increase in risk of developing dementia (Tang et al., 2021). Overall, people with established AD have lower triiodothyronine (T3), but no difference in TSH or thyroxine (T4) compared to controls (Dolatshahi et al., 2023), while a central pattern of hypothyroidism (reduced TRH, TSH, T3 and T4) may be seen in people with advanced AD (Chen et al., 2013; Yong-Hong et al., 2013).

The new automatic segmentation pipelines will be useful for future studies to determine whether the abnormalities seen in the HPA, HPG, the hypothalamic-pituitary-thyroid and the hypothalamic-pituitary-somatotropic (growth hormone) axes in people with AD are related to hypothalamic pathology. This may have important implications for new treatment targets and to understand the increased prevalence of osteoporosis in people with AD (Xie et al., 2023). On study has found that bone mineral density was associated with hypothalamic volume in AD, but not in healthy controls (Loskutova et al., 2010). Many of the hormones under hypothalamic control influence bone homeostasis, as seen by the prevalence of osteoporosis in people with early menopause, and in conditions such as hyperprolactinaemia, hyperthyroidism and hypocortisolaemia (Niwczyk et al., 2023). In addition to the hormonal route, neuronal signalling from the hypothalamic nuclei also act directly on osteoblasts through their innervation by the sympathetic nervous system (Niwczyk et al., 2023). Understanding the causes of osteoporosis in AD is important due to the significantly higher risk of bone fractures, which in turn is associated with a significant morbidity and mortality.

### 4.4 Limitations

Most of the studies included in this review were small case-control studies, introducing possible bias, and limiting the generalisability of the conclusions to the general population of people with AD. The meta-analysis had significant between-study heterogeneity, which is likely a reflection of the variation in the study population size, MRI acquisition protocol and analysis protocol. The inclusion of people with different stages of AD may also have contributed to the heterogeneity, although the range of cognitive scores (MMSE range: 19.2-26.1) suggests that most people had mild-to-moderate dementia. Despite the heterogeneity in study design, only two studies (Ahmed et al., 2015; Kim et al., 2016) did not find a significant difference in hypothalamic volume, while no study reported a larger hypothalamus in AD. While there was no suggestion of publication bias, the small number of included studies which could be included in the meta-analysis may have made the funnel plot difficult to interpret. Finally, due to the absence of longitudinal studies, or studies using AD biomarkers, it is not possible to make any firm conclusions about the cause of a smaller hypothalamus, or the changes in volume across the AD disease process.

## 5.0 Conclusion

In this meta-analysis, we found evidence that the hypothalamus is smaller in people with AD (-10.1%) compared to healthy controls, as shown by *in vivo* MRI structural imagining. One study also found qualitative evidence that the hypothalamus is reduced in DLB, but further studies are required to confirm this finding. Hypothalamic volume is also smaller in other neurodegenerative conditions, in particular FTD and ALS, suggesting that the hypothalamus may be vulnerable to neurodegeneration in the context of many different pathological processes. Hypothalamic volume loss may be greater in men than women, and the arcuate nuclei may be affected less than others. Finally, a smaller hypothalamus may be associated with plasma levels of sex hormones and reduced bone mineral density in AD, indicating that hypothalamic involvement may have specific functional consequences. Further studies will need to assess how hypothalamic pathology evolves during the disease process in AD and LBD and its impact on weight, sleep, endocrine function and clinically meaningful symptomatology.

## 8.0 Data availability

Data is available from the corresponding author upon reasonable request.

## References

Ahmad, M.H., Fatima, M., Mondal, A.C., 2019. Role of hypothalamic-pituitary-adrenal axis, hypothalamic-pituitary-gonadal axis and insulin signaling in the pathophysiology of Alzheimer’s disease. Neuropsychobiology. 77, 197–205. 10.1159/000495521

Ahmed, R., Latheef, S., Bartley, L., Irish, M., Halliday, G., Kiernan, M., Hodges, J., Piguet, O., 2015. Eating behavior in frontotemporal dementia: peripheral hormones vs hypothalamic pathology. Neurology. 85, 1310–1317. 10.1212/WNL.0000000000002018

Baker, L.D., Barsness, S.M., Borson, S., Merriam, G.R., Friedman, S.D., Craft, S., Vitiello, M.V., 2012. Effects of growth hormone–releasing hormone on cognitive function in adults with mild cognitive impairment and healthy older adults. Arch. Neurol. 69, 1420–1429. 10.1001/archneurol.2012.1970

Baloyannis, S.J., Mavroudis, I., Mitilineos, D., Baloyannis, I.S., Costa, V.G., 2015. The hypothalamus in Alzheimer’s disease: a golgi and electron microscope study. Am. J. Alzheimers. Dis. Other. Demen. 30, 478–487. 10.1177/1533317514556876

Bang, J., Spina, S., Miller, B.L., 2015. Frontotemporal dementia. Lancet. 386, 1672–1682. 10.1016/S0140-6736(15)00461-4

Baron, J.C., Chételat, G., Desgranges, B., Perchey, G., Landeau, B., de la Sayette, V., Eustache, F., 2001. In vivo mapping of gray matter loss with voxel-based morphometry in mild Alzheimer’s disease. Neuroimage. 14, 298–309. 10.1006/nimg.2001.0848

Benarroch, E.E., Schmeichel, A.M., Parisi, J.E., Low, P.A., 2015. Histaminergic tuberomammillary neuron loss in multiple system atrophy and dementia with Lewy bodies. Mov. Disord. 30, 1133–1139. 10.1002/mds.26287

Billot, B., Bocchetta, M., Todd, E., Dalca, A.V., Rohrer, J.D., Iglesias, J.E., 2020. Automated segmentation of the hypothalamus and associated subunits in brain MRI. Neuroimage. 223, 117287. 10.1016/j.neuroimage.2020.117287

Bocchetta, M., Gordon, E., Manning, E., Barnes, J., Cash, D.M., Espak, M., Thomas, D.L., Modat, M., Rossor, M.N., Warren, J.D., Ourselin, S., Frisoni, G.B., Rohrer, J.D., 2015. Detailed volumetric analysis of the hypothalamus in behavioral variant frontotemporal dementia. J. Neurol. 262, 2635–2642. 10.1007/s00415-015-7885-2

Bowen, R.L., Isley, J.P., Atkinson, R.L., 2000. An association of elevated serum gonadotropin concentrations and Alzheimer disease? J. Neuroendocrinol. 12, 351–354. 10.1046/j.1365-2826.2000.00461.x

Braak, H., Braak, E., 1991. Neuropathological stageing of Alzheimer-related changes. Acta. Neuropathol. 82, 239–259. 10.1007/BF00308809

Braak, H., Del Tredici, K., Rüb, U., de Vos, R.A.I., Jansen Steur, E.N.H., Braak, E., 2003. Staging of brain pathology related to sporadic Parkinson’s disease. Neurobiol. Aging. 24, 197–211. 10.1016/s0197-4580(02)00065-9

Butchart, J., Birch, B., Bassily, R., Wolfe, L., Holmes, C., 2013. Male sex hormones and systemic inflammation in Alzheimer disease. Alzheimer. Dis. Assoc. Disord. 27, 153–156. 10.1097/WAD.0b013e318258cd63

Callen, D., Black, S., Caldwell, C., 2002. Limbic system perfusion in Alzheimer’s disease measured by MRI-coregistered HMPAO SPET. Eur. J. Nucl. Med. Mol. Imaging. 29, 899–906. 10.1007/s00259-002-0816-3

Callen, D., Black, S., Caldwell, C., Grady, C., 2004. The influence of sex on limbic volume and perfusion in AD. Neurobiol. Aging. 25, 761–770. 10.1016/j.neurobiolaging.2003.08.011

Callen, D., Black, S., Gao, F., Caldwell, C., Szalai, J., 2001. Beyond the hippocampus: MRI volumetry confirms widespread limbic atrophy in AD. Neurology. 57, 1669–1674. 10.1212/wnl.57.9.1669

Chang, J., Shaw, T.B., Holdom, C.J., McCombe, P.A., Henderson, R.D., Fripp, J., Barth, M., Guo, C.C., Ngo, S.T., Steyn, F.J., Alzheimer’s Disease Neuroimaging Initiative, 2023. Lower hypothalamic volume with lower body mass index is associated with shorter survival in patients with amyotrophic lateral sclerosis. Eur. J. Neurol. 30, 57–68. 10.1111/ene.15589

Chang, J., Steyn, F., Ngo, S., Henderson, R., Guo, C., Bollmann, S., Fripp, J., Barth, M., Shaw, T., 2022. Open-source hypothalamic-forniX (OSHy-X) atlases and segmentation tool for 3T and 7T. Journal of Open Source Software. 7, 4368. 10.21105/joss.04368

Chen, J.-M., Huang, C.-Q., Ai, M., Kuang, L., 2013. Circadian rhythm of TSH levels in subjects with Alzheimer’s disease (AD). Aging. Clin. Exp. Res. 25, 153–157. 10.1007/s40520-013-0025-x

Coll, A.P., Farooqi, I.S., O’Rahilly, S., 2007. The hormonal control of food intake. Cell. 129, 251–262. 10.1016/j.cell.2007.04.001

Compta, Y., Santamaria, J., Ratti, L., Tolosa, E., Iranzo, A., Muñoz, E., Valldeoriola, F., Casamitjana, R., Ríos, J., Marti, M.J., 2009. Cerebrospinal hypocretin, daytime sleepiness and sleep architecture in Parkinson’s disease dementia. Brain. 132, 3308–3317. 10.1093/brain/awp263

Copenhaver, B.R., Rabin, L.A., Saykin, A.J., Roth, R.M., Wishart, H.A., Flashman, L.A., Santulli, R.B., McHugh, T.L., Mamourian, A.C., 2006. The fornix and mammillary bodies in older adults with Alzheimer’s disease, mild cognitive impairment, and cognitive complaints: a volumetric MRI study. Psychiatry. Res. 147, 93–103. 10.1016/j.pscychresns.2006.01.015

Cross, D.J., Anzai, Y., Petrie, E.C., Martin, N., Richards, T.L., Maravilla, K.R., Peskind, E.R., Minoshima, S., 2013. Loss of olfactory tract integrity affects cortical metabolism in the brain and olfactory regions in aging and mild cognitive impairment. J. Nucl. Med. 54, 1278–1284. 10.2967/jnumed.112.116558

Csernansky, J.G., Dong, H., Fagan, A.M., Wang, L., Xiong, C., Holtzman, D.M., Morris, J.C., 2006. Plasma cortisol and progression of dementia in subjects with Alzheimer-type dementia. Am. J. Psychiatry. 163, 2164–2169. 10.1176/ajp.2006.163.12.2164

Desai, K., McManus, J.M., Sharifi, N., 2021. Hormonal therapy for prostate cancer. Endocr. Rev. 42, 354–373. 10.1210/endrev/bnab002

Dolatshahi, M., Salehipour, A., Saghazadeh, A., Sanjeari Moghaddam, H., Aghamollaii, V., Fotouhi, A., Tafakhori, A., 2023. Thyroid hormone levels in Alzheimer disease: a systematic review and meta-analysis. Endocrine. 79, 252–272. 10.1007/s12020-022-03190-w

Elgh, E., Lindqvist Astot, A., Fagerlund, M., Eriksson, S., Olsson, T., Näsman, B., 2006. Cognitive dysfunction, hippocampal atrophy and glucocorticoid feedback in Alzheimer’s disease. Biol. Psychiatry. 59, 155–161. 10.1016/j.biopsych.2005.06.017

Fronczek, R., van Geest, S., Frölich, M., Overeem, S., Roelandse, F.W.C., Lammers, G.J., Swaab, D.F., 2012. Hypocretin (orexin) loss in Alzheimer’s disease. Neurobiol. Aging. 33, 1642–1650. 10.1016/j.neurobiolaging.2011.03.014

Gabelle, A., Jaussent, I., Hirtz, C., Vialaret, J., Navucet, S., Grasselli, C., Robert, P., Lehmann, S., Dauvilliers, Y., 2017. Cerebrospinal fluid levels of orexin-A and histamine, and sleep profile within the Alzheimer process. Neurobiol. Aging. 53, 59–66. 10.1016/j.neurobiolaging.2017.01.011

Gabery, S., Georgiou-Karistianis, N., Lundh, S.H., Cheong, R.Y., Churchyard, A., Chua, P., Stout, J.C., Egan, G.F., Kirik, D., Petersén, Å., 2015. Volumetric analysis of the hypothalamus in Huntington disease using 3T MRI: the IMAGE-HD study. PLoS One. 10, e0117593. 10.1371/journal.pone.0117593

Goldstein, J.M., Seidman, L.J., Horton, N.J., Makris, N., Kennedy, D.N., Caviness, V.S., Faraone, S.V., Tsuang, M.T., 2001. Normal sexual dimorphism of the adult human brain assessed by in vivo magnetic resonance imaging. Cereb. Cortex. 11, 490–497. 10.1093/cercor/11.6.490

Goodman, R.A., Lochner, K.A., Thambisetty, M., Wingo, T.S., Posner, S.F., Ling, S.M., 2017. Prevalence of dementia subtypes in United States Medicare fee-for-service beneficiaries, 2011–2013. Alzheimers Dement. 13, 28–37. 10.1016/j.jalz.2016.04.002

Gorges, M., Vercruysse, P., Müller, H.-P., Huppertz, H.-J., Rosenbohm, A., Nagel, G., Weydt, P., Petersén, Å., Ludolph, A.C., Kassubek, J., Dupuis, L., 2017. Hypothalamic atrophy is related to body mass index and age at onset in amyotrophic lateral sclerosis. J. Neurol. Neurosurg. Psychiatry. 88, 1033–1041. 10.1136/jnnp-2017-315795

Grundman, M., Corey-Bloom, J., Jernigan, T., Archibald, S., Thal, L., 1996. Low body weight in Alzheimer’s disease is associated with mesial temporal cortex atrophy. Neurology. 46, 1585–1591. 10.1212/wnl.46.6.1585

Guo, J., Jiang, Z., Biswal, B.B., Zhou, B., Xie, D., Gao, Q., Sheng, W., Chen, Hui, Zhang, Y., Fan, Y., Wang, J., Liu, C., Chen, H., 2022. Hypothalamic atrophy, expanded CAG repeat, and low body mass index in spinocerebellar ataxia type 3. Mov. Disord. 37, 1541–1546. 10.1002/mds.29029

Hall, A.M., Moore, R.Y., Lopez, O.L., Kuller, L., Becker, J.T., 2008. Basal forebrain atrophy is a presymptomatic marker for Alzheimer’s disease. Alzheimers. Dement. 4, 271–279. 10.1016/j.jalz.2008.04.005

Harper, D.G., Stopa, E.G., Kuo-Leblanc, V., McKee, A.C., Asayama, K., Volicer, L., Kowall, N., Satlin, A., 2008. Dorsomedial SCN neuronal subpopulations subserve different functions in human dementia. Brain. 131, 1609–1617. 10.1093/brain/awn049

Herring, W.J., Ceesay, P., Snyder, E., Bliwise, D., Budd, K., Hutzelmann, J., Stevens, J., Lines, C., Michelson, D., 2020. Polysomnographic assessment of suvorexant in patients with probable Alzheimer’s disease dementia and insomnia: a randomized trial. Alzheimers. Dement. 16, 541–551. 10.1002/alz.12035

Higgins, J.P.T., Thompson, S.G., 2002. Quantifying heterogeneity in a meta-analysis. Stat. Med. 21, 1539–1558. 10.1002/sim.1186

Huang, C.-W., Lui, C.-C., Chang, W.-N., Lu, C.-H., Wang, Y.-L., Chang, C.-C., 2009. Elevated basal cortisol level predicts lower hippocampal volume and cognitive decline in Alzheimer’s disease. J. Clin. Neurosci. 16, 1283–1286. 10.1016/j.jocn.2008.12.026

Ishii, M., Iadecola, C., 2015. Metabolic and non-cognitive manifestations of Alzheimer’s disease: the hypothalamus as both culprit and target of pathology. Cell. Metab. 22, 761–776. 10.1016/j.cmet.2015.08.016

Ismail, Z., Herrmann, N., Rothenburg, L., Cotter, A., Leibovitch, F., Rafi-Tari, S., Black, S., Lanctot, K., 2008. A functional neuroimaging study of appetite loss in Alzheimer’s disease. J. Neurol. Sci. 271, 97–103. 10.1016/j.jns.2008.03.023

Joling, M., Vriend, C., Raijmakers, P.G.H.M., van der Zande, J.J., Lemstra, A.W., Berendse, H.W., Booij, J., van den Heuvel, O.A., 2019. Striatal DAT and extrastriatal SERT binding in early-stage Parkinson’s disease and dementia with Lewy bodies, compared with healthy controls: an 123I-FP-CIT SPECT study. Neuroimage. Clin. 22, 101755. 10.1016/j.nicl.2019.101755

Kasanuki, K., Iseki, E., Kondo, D., Fujishiro, H., Minegishi, M., Sato, K., Katsuse, O., Hino, H., Kosaka, K., Arai, H., 2014. Neuropathological investigation of hypocretin expression in brains of dementia with Lewy bodies. Neurosci. Lett. 569, 68–73. 10.1016/j.neulet.2014.03.020

Kim, H.-J., Im, H., Kim, J., Han, J.-Y., De Leon, M., Deshpande, A., Moon, W.-J., 2016. Brain atrophy of secondary REM-sleep behavior disorder in neurodegenerative disease. J. Alzheimers. Dis. 52, 1101–1109. 10.3233/JAD-151197

Koren, T., Fisher, E., Webster, L., Livingston, G., Rapaport, P., 2023. Prevalence of sleep disturbances in people with dementia living in the community: a systematic review and meta-analysis. Ageing. Res. Rev. 83, 101782. 10.1016/j.arr.2022.101782

Lanctot, K., Herrmann, N., Nadkarni, N., Leibovitch, F., Caldwell, C., Black, S., 2004. Medial Temporal Hypoperfusion and Aggression in Alzheimer Disease. Arch. Neurol. 61, 1731–1737. 10.1001/archneur.61.11.1731

Lanctot, K., Moosa, S., Herrmann, N., Leibovitch, F., Rothenburg, L., Cotter, A., Black, S., 2007. A SPECT study of apathy in Alzheimer’s disease. Dement. Geriatr. Cogn. Disord. 24, 65–72. 10.1159/000103633

Liguori, C., Chiaravalloti, A., Nuccetelli, M., Izzi, F., Sancesario, G., Cimini, A., Bernardini, S., Schillaci, O., Mercuri, N., Fabio, P., 2017. Hypothalamic dysfunction is related to sleep impairment and CSF biomarkers in Alzheimer’s disease. J. Neurol. 264, 2215–2223. 10.1007/s00415-017-8613-x

Liguori, C., Nuccetelli, M., Izzi, F., Sancesario, G., Romigi, A., Martorana, A., Amoroso, C., Bernardini, S., Marciani, M.G., Mercuri, N.B., Placidi, F., 2016. Rapid eye movement sleep disruption and sleep fragmentation are associated with increased orexin-A cerebrospinal-fluid levels in mild cognitive impairment due to Alzheimer’s disease. Neurobiol. Aging. 40, 120–126. 10.1016/j.neurobiolaging.2016.01.007

Liguori, C., Romigi, A., Nuccetelli, M., Zannino, S., Sancesario, G., Martorana, A., Albanese, M., Mercuri, N.B., Izzi, F., Bernardini, S., Nitti, A., Sancesario, G.M., Sica, F., Marciani, M.G., Placidi, F., 2014. Orexinergic system dysregulation, sleep impairment, and cognitive decline in Alzheimer disease. JAMA. Neurol. 71, 1498–1505. 10.1001/jamaneurol.2014.2510

Lim, A.S.P., Ellison, B.A., Wang, J.L., Yu, L., Schneider, J.A., Buchman, A.S., Bennett, D.A., Saper, C.B., 2014. Sleep is related to neuron numbers in the ventrolateral preoptic/intermediate nucleus in older adults with and without Alzheimer’s disease. Brain. 137, 2847–2861. 10.1093/brain/awu222

Liu, S., Ren, Q., Gong, G., Sun, Y., Zhao, B., Ma, X., Zhang, N., Zhong, S., Lin, Y., Wang, W., Zheng, R., Yu, X., Yun, Y., Zhang, D., Shao, K., Lin, P., Yuan, Y., Dai, T., Zhang, Y., Li, L., Li, W., Zhao, Y., Shan, P., Meng, X., Yan, C., 2022. Hypothalamic subregion abnormalities are related to body mass index in patients with sporadic amyotrophic lateral sclerosis. J. Neurol. 269, 2980–2988. 10.1007/s00415-021-10900-3

Livingston, G., Huntley, J., Liu, K.Y., Costafreda, S.G., Selbæk, G., Alladi, S., Ames, D., Banerjee, S., Burns, A., Brayne, C., Fox, N.C., Ferri, C.P., Gitlin, L.N., Howard, R., Kales, H.C., Kivimäki, M., Larson, E.B., Nakasujja, N., Rockwood, K., Samus, Q., Shirai, K., Singh-Manoux, A., Schneider, L.S., Walsh, S., Yao, Y., Sommerlad, A., Mukadam, N., 2024. Dementia prevention, intervention, and care: 2024 report of the Lancet standing Commission. Lancet. 404, 572–628. 10.1016/S0140-6736(24)01296-0

Loskutova, N., Honea, R., Brooks, W., Burns, J., 2010. Reduced limbic and hypothalamic volumes correlate with bone density in early Alzheimer’s disease. J Alzheimers. Dis. 20, 313–322. 10.3233/JAD-2010-1364

Lucey, B.P., Liu, H., Toedebusch, C.D., Freund, D., Redrick, T., Chahin, S.L., Mawuenyega, K.G., Bollinger, J.G., Ovod, V., Barthélemy, N.R., Bateman, R.J., 2023. Suvorexant acutely decreases tau phosphorylation and Aβ in the human CNS. Ann. Neurol. 94, 27–40. 10.1002/ana.26641

Nerattini, M., Rubino, F., Jett, S., Andy, C., Boneu, C., Zarate, C., Carlton, C., Loeb-Zeitlin, S., Havryliuk, Y., Pahlajani, S., Williams, S., Berti, V., Christos, P., Fink, M., Dyke, J.P., Brinton, R.D., Mosconi, L., 2023. Elevated gonadotropin levels are associated with increased biomarker risk of Alzheimer’s disease in midlife women. Front. Dement. 2, 1303256. 10.3389/frdem.2023.1303256

Nestor, P.J., Fryer, T.D., Smielewski, P., Hodges, J.R., 2003. Limbic hypometabolism in Alzheimer’s disease and mild cognitive impairment. Ann. Neurol. 54, 343–351. 10.1002/ana.10669

Niwczyk, O., Grymowicz, M., Szczęsnowicz, A., Hajbos, M., Kostrzak, A., Budzik, M., Maciejewska-Jeske, M., Bala, G., Smolarczyk, R., Męczekalski, B., 2023. Bones and hormones: interaction between hormones of the hypothalamus, pituitary, adipose tissue and bone. Int. J. Mol. Sci. 24, 6840. 10.3390/ijms24076840

O’Brien, J., Ames, D., Schweitzer, I., Colman, P., Desmond, P., Tress, B., 1996. Clinical and magnetic resonance imaging correlates of hypothalamic-pituitary-adrenal axis function in depression and Alzheimer’s disease. Br. J. Psychiatry. 168, 679–687. 10.1192/bjp.168.6.679

Ofori, E., Solis, A., Punjani, N., 2024. The association among hypothalamic subnits, gonadotropic and sex hormone plasmas levels in Alzheimer’s disease. Brain Sci. 14, 276. 10.3390/brainsci14030276

Page, M.J., McKenzie, J.E., Bossuyt, P.M., Boutron, I., Hoffmann, T.C., Mulrow, C.D., Shamseer, L., Tetzlaff, J.M., Akl, E.A., Brennan, S.E., Chou, R., Glanville, J., Grimshaw, J.M., Hróbjartsson, A., Lalu, M.M., Li, T., Loder, E.W., Mayo-Wilson, E., McDonald, S., McGuinness, L.A., Stewart, L.A., Thomas, J., Tricco, A.C., Welch, V.A., Whiting, P., Moher, D., 2021. The PRISMA 2020 statement: an updated guideline for reporting systematic reviews. BMJ. 372, n71. 10.1136/bmj.n71

Piguet, O., Petersén, A., Yin Ka Lam, B., Gabery, S., Murphy, K., Hodges, J.R., Halliday, G.M., 2011. Eating and hypothalamus changes in behavioral-variant frontotemporal dementia. Ann. Neurol. 69, 312–319. 10.1002/ana.22244

Popp, J., Wolfsgruber, S., Heuser, I., Peters, O., Hüll, M., Schröder, J., Möller, H.-J., Lewczuk, P., Schneider, A., Jahn, H., Luckhaus, C., Perneczky, R., Frölich, L., Wagner, M., Maier, W., Wiltfang, J., Kornhuber, J., Jessen, F., 2015. Cerebrospinal fluid cortisol and clinical disease progression in MCI and dementia of Alzheimer’s type. Neurobiol. Aging. 36, 601–607. 10.1016/j.neurobiolaging.2014.10.031

Purba, J.S., Hofman, M.A., Swaab, D.F., 1994. Decreased number of oxytocin-immunoreactive neurons in the paraventricular nucleus of the hypothalamus in Parkinson’s disease. Neurology. 44, 84–89. 10.1212/wnl.44.1.84

Raadsheer, F.C., van Heerikhuize, J.J., Lucassen, P.J., Hoogendijk, W.J., Tilders, F.J., Swaab, D.F., 1995. Corticotropin-releasing hormone mRNA levels in the paraventricular nucleus of patients with Alzheimer’s disease and depression. Am. J. Psychiatry. 152, 1372–1376. 10.1176/ajp.152.9.1372

Saper, C.B., German, D.C., 1987. Hypothalamic pathology in Alzheimer’s disease. Neurosci. Lett. 74, 364–370. 10.1016/0304-3940(87)90325-9

Saper, C.B., Scammell, T.E., Lu, J., 2005. Hypothalamic regulation of sleep and circadian rhythms. Nature. 437, 1257–1263. 10.1038/nature04284

Schultz, C., Braak, H., Braak, E., 1996. A sex difference in neurodegeneration of the human hypothalamus. Neurosci. Lett. 212, 103–106. 10.1016/0304-3940(96)12787-7

Schultz, C., Ghebremedhin, E., Braak, E., Braak, H., 1999. Sex-dependent cytoskeletal changes of the human hypothalamus develop independently of Alzheimer’s disease. Exp. Neurol. 160, 186–193. 10.1006/exnr.1999.7185

Shan, L., Bossers, K., Unmehopa, U., Bao, A.-M., Swaab, D.F., 2012. Alterations in the histaminergic system in Alzheimer’s disease: a postmortem study. Neurobiol. Aging. 33, 2585–2598. 10.1016/j.neurobiolaging.2011.12.026

Shapiro, N.L., Todd, E.G., Billot, B., Cash, D.M., Iglesias, J.E., Warren, J.D., Rohrer, J.D., Bocchetta, M., 2022. In vivo hypothalamic regional volumetry across the frontotemporal dementia spectrum. Neuroimage. Clin. 35, 103084. 10.1016/j.nicl.2022.103084

Smith, M.A., Bowen, R.L., Nguyen, R.Q., Perry, G., Atwood, C.S., Rimm, A.A., 2018. Putative gonadotropin-releasing hormone agonist therapy and dementia: an application of Medicare hospitalization claims data. J. Alzheimers. Dis. 63, 1269–1277. 10.3233/JAD-170847

Soneson, C., Fontes, M., Zhou, Y., Denisov, V., Paulsen, J.S., Kirik, D., Petersén, A., Huntington Study Group PREDICT-HD investigators, 2010. Early changes in the hypothalamic region in prodromal Huntington disease revealed by MRI analysis. Neurobiol. Dis. 40, 531–543. 10.1016/j.nbd.2010.07.013

Soysal, P., Tan, S.G., Rogowska, M., Jawad, S., Smith, L., Veronese, N., Tsiptsios, D., Tsamakis, K., Stewart, R., Mueller, C., 2021. Weight loss in Alzheimer’s disease, vascular dementia and dementia with Lewy bodies: impact on mortality and hospitalization by dementia subtype. Int. J. Geriatr. Psychiatry. 37. 10.1002/gps.5659

Tang, X., Song, Z.-H., Wang, D., Yang, J., Augusto Cardoso, M., Zhou, J.-B., Simó, R., 2021. Spectrum of thyroid dysfunction and dementia: a dose-response meta-analysis of 344,248 individuals from cohort studies. Endocr. Connect. 10, 410–421. 10.1530/EC-21-0047

Toledo, J.B., Toledo, E., Weiner, M.W., Jack, C.R., Jagust, W., Lee, V.M.Y., Shaw, L.M., Trojanowski, J.Q., Alzheimer’s Disease Neuroimaging Initiative, 2012. Cardiovascular risk factors, cortisol, and amyloid-β deposition in Alzheimer’s disease neuroimaging initiative. Alzheimers. Dement. 8, 483–489. 10.1016/j.jalz.2011.08.008

Tse, N.Y., Bocchetta, M., Todd, E.G., Devenney, E.M., Tu, S., Caga, J., Hodges, J.R., Halliday, G.M., Irish, M., Kiernan, M.C., Piguet, O., Rohrer, J.D., Ahmed, R.M., 2023. Distinct hypothalamic involvement in the amyotrophic lateral sclerosis-frontotemporal dementia spectrum. Neuroimage. Clin. 37, 103281. 10.1016/j.nicl.2022.103281

Wells, G., Shea, B., O’Connell, D., Peterson, J., Welch, V., Losos, M., Tugwell, P., 2021. The Newcastle-Ottawa Scale (NOS) for assessing the quality of nonrandomised studies in meta-analyses. https://www.ohri.ca/programs/clinical_epidemiology/oxford.asp (accessed 3rd Sept 2024).

Westwood, A.J., Beiser, A., DeCarli, C., Harris, T.B., Chen, T.C., He, X., Roubenoff, R., Pikula, A., Au, R., Braverman, L.E., Wolf, P.A., Vasan, R.S., Seshadri, S., 2014. Insulin-like growth factor-1 and risk of Alzheimer dementia and brain atrophy. Neurology. 82, 1613–1619. 10.1212/WNL.0000000000000382

Whitwell, J.L., Weigand, S.D., Shiung, M.M., Boeve, B.F., Ferman, T.J., Smith, G.E., Knopman, D.S., Petersen, R.C., Benarroch, E.E., Josephs, K.A., Jack, C.R., Jr, 2007. Focal atrophy in dementia with Lewy bodies on MRI: a distinct pattern from Alzheimer’s disease. Brain. 130, 708–719. 10.1093/brain/awl388

Woolley, J.D., Khan, B.K., Natesan, A., Karydas, A., Dallman, M., Havel, P., Miller, B.L., Rankin, K.P., 2014. Satiety-related hormonal dysregulation in behavioral variant frontotemporal dementia. Neurology. 82, 512–520. 10.1212/WNL.0000000000000106

Xie, C., Wang, C., Luo, H., 2023. Increased risk of osteoporosis in patients with cognitive impairment: a systematic review and meta-analysis. BMC. Geriatr. 23, 1–10. 10.1186/s12877-023-04548-z

Yasui, K., Inoue, Y., Kanbayashi, T., Nomura, T., Kusumi, M., Nakashima, K., 2006. CSF orexin levels of Parkinson’s disease, dementia with Lewy bodies, progressive supranuclear palsy and corticobasal degeneration. J. Neurol. Sci. 250, 120–123. 10.1016/j.jns.2006.08.004

Yong-Hong, L., Xiao-Dong, P., Chang-Quan, H., Bo, Y., Qing-Xiu, L., 2013. Hypothalamic-pituitary-thyroid axis in patients with Alzheimer disease (AD). J. Investig. Med. 61, 578–581. 10.2310/JIM.0b013e318280aafb

Zhou, H.-H., Yu, Z., Luo, L., Xie, F., Wang, Y., Wan, Z., 2021. The effect of hormone replacement therapy on cognitive function in healthy postmenopausal women: a meta-analysis of 23 randomized controlled trials. Psychogeriatrics. 21, 926–938. 10.1111/psyg.12768

