## Supplemental Table 1 for "Hypothalamic structure and function in Alzheimer’s disease and Lewy-body dementia: a systematic review and meta-analysis"

**Table S1.** List of papers excluded after full-text review.

| **First author** | **Title** | **Reason for exclusion** |
| --- | --- | --- |
| Whitwell, et al. | Patterns of Atrophy on MRI in Dementia with Lewy Bodies and Alzheimer's Disease: A Voxel-Based Morphometry Study: P02.116. | Full text not available. |
| Callen et al. | The Limbic System in Alzheimer's Disease: A Volumetric Study Using Magnetic Resonance Imaging. | Full text not available. |
| Yang et al. | Altered cortical and subcortical morphometric features and asymmetries in the subjective cognitive decline and mild cognitive impairment | Subjects without dementia. |
| Liang et al. | Altered directional connectivity between emotion network and motor network in Parkinson's disease with depression. | Subjects without dementia. |
| van Mierlo et al. | Depressive Symptoms in Parkinson's Disease Are Related To Decreased Hippocampus and Amygdala Volume. | Subjects without dementia. |
| Brown et al. | Development, validation and application of a new fornix template for studies of aging and preclinical Alzheimer's disease. | Subjects without dementia. |
| Stankeviciute et al. | Differential effects of sleep on brain structure and metabolism at the preclinical stages of AD. | Subjects without dementia. |
| Dayan et al. | Disrupted hypothalamic functional connectivity in patients with PD and autonomic dysfunction. | Subjects without dementia. |
| Politis et al. | Evidence of dopamine dysfunction in the hypothalamus of patients with Parkinson's disease: An in vivo 11C-raclopride PET study. | Subjects without dementia. |
| Breen et al. | Hypothalamic Volume Loss Is Associated With Reduced Melatonin Output in Parkinson's Disease. | Subjects without dementia. |
| Pavese et al. | In vivo assessment of brain monoamine systems in parkin gene carriers: A PET study. | Subjects without dementia. |
| Niccolini et al. | Loss of phosphodiesterase 4 in Parkinson disease: Relevance to cognitive deficits. | Subjects without dementia. |
| Gorges et al. | Morphological MRI investigations of the hypothalamus in 232 individuals with Parkinson's disease. | Subjects without dementia. |
| Lebedeva et al. | MRI-based classification models in prediction of mild cognitive impairment and dementia in late-life depression | Subjects without dementia. |
| Schulz et al. | Nucleus basalis of Meynert degeneration precedes and predicts cognitive impairment in Parkinson's disease. | Subjects without dementia. |
| Asano et al. | Reduced gray matter volume in the default-mode network associated with insulin resistance | Subjects without dementia. |
| Wile et al. | Serotonin and dopamine transporter PET changes in the premotor phase of LRRK2 parkinsonism: cross-sectional studies | Subjects without dementia. |
| van der Velpen et al. | Subcortical brain structures and the risk of dementia in the Rotterdam Study. | Subjects without dementia. |
| Skoog et al. | A Population-Based Study on Blood Pressure and Brain Atrophy in 85-Year-Olds. | No hypothalamic imaging. |
| Junque et al. | Amygdalar and Hippocampal Mri Volumetric Reductions in Parkinson's Disease With Dementia. | No hypothalamic imaging. |
| Magri et al. | Association between changes in adrenal secretion and cerebral morphometric correlates in normal aging and senile dementia | No hypothalamic imaging. |
| Rozalem Aranha et al. | Basal forebrain atrophy along the Alzheimer's disease continuum in adults with Down syndrome. | No hypothalamic imaging. |
| Fernandez-Cabello et al. | Basal forebrain volume reliably predicts the cortical spread of Alzheimer's degeneration. | No hypothalamic imaging. |
| Loskutova et al. | Bone density and brain atrophy in early Alzheimer's disease | No hypothalamic imaging. |
| O'Brien et al. | Clinical and Magnetic Resonance Imaging Correlates of Hypothalamic-Pituitary-Adrenal Axis Function in Depression and Alzheimer's Disease. | No hypothalamic imaging. |
| Elgh et al. | Cognitive dysfunction, hippocampal atrophy and glucocorticoid feedback in Alzheimer's disease | No hypothalamic imaging. |
| Ojkowska et al. | Correlations Between Cerebellar and Brain Volumes, Cognitive Impairments, ApoE Levels, and APOE Genotypes in Patients with AD and MCI. | No hypothalamic imaging. |
| Li et al. | Early-stage differentiation between Alzheimer's disease and frontotemporal lobe degeneration: Clinical, neuropsychology, and neuroimaging features | No hypothalamic imaging. |
| Goldman et al. | Entorhinal cortex atrophy differentiates Parkinson's disease patients with and without dementia. | No hypothalamic imaging. |
| Hoefer et al. | Fear conditioning in frontotemporal lobar degeneration and Alzheimer's disease. | No hypothalamic imaging. |
| Saygin et al. | High-resolution magnetic resonance imaging reveals nuclei of the human amygdala: manual segmentation to automatic atlas. | No hypothalamic imaging. |
| Sabattoli et al. | Hippocampal shape differences in dementia with Lewy bodies. | No hypothalamic imaging. |
| Plachti et al. | Hippocampus co-atrophy pattern in dementia deviates from covariance patterns across the lifespan. | No hypothalamic imaging. |
| Mazere et al. | In vivo SPECT imaging of vesicular acetylcholine transporter using [123I]-IBVM in early Alzheimer's disease. | No hypothalamic imaging. |
| Cavedo et al. | Local amygdala structural differences with 3T MRI in patients with Alzheimer disease. | No hypothalamic imaging. |
| Prasher et al. | Magnetic resonance imaging, Down's syndrome and Alzheimer's disease: research and clinical implications. | No hypothalamic imaging. |
| Dhikav et al. | Medial temporal lobe atrophy in Alzheimer's disease/mild cognitive impairment with depression. | No hypothalamic imaging. |
| Zarow et al. | MRI shows more severe hippocampal atrophy and shape deformation in hippocampal sclerosis than in Alzheimer's disease | No hypothalamic imaging. |
| Stout et al. | Regional Cerebral Volume Loss Associated With Verbal Learning and Memory in Dementia of the Alzheimer Type. | No hypothalamic imaging. |
| Blautzik et al. | Relationship Between Body Mass Index, ApoE4 Status, and PET-Based Amyloid and Neurodegeneration Markers in Amyloid-Positive Subjects with Normal Cognition or Mild Cognitive Impairment. | No hypothalamic imaging. |
| Mueller et al. | Selective effect of Apo e4 on CA3 and dentate in normal aging and Alzheimer's disease using high resolution MRI at 4 T. | No hypothalamic imaging. |
| Skup et al. | Sex differences in grey matter atrophy patterns among AD and aMCI patients: Results from ADNI. | No hypothalamic imaging. |
| Ebmeier et al. | Temporal lobe abnormalities in dementia and depression: a study using high resolution single photon emission tomography and magnetic resonance imaging. | No hypothalamic imaging. |
| Liu et al. | The abnormal functional connectivity between the hypothalamus and the temporal gyrus underlying depression in Alzheimer's disease patients | No hypothalamic imaging. |
| Chen et al. | Voxel-level comparison of arterial spin-labeled perfusion MRI and FDG-PET in Alzheimer disease. | No hypothalamic imaging. |
| Barber et al. | White matter lesions on magnetic resonance imaging in dementia with Lewy bodies, Alzheimer's disease, vascular dementia, and normal aging | No hypothalamic imaging. |
| Liguori et al. | Hypothalamic dysfunction is related to sleep impairment and CSF biomarkers in Alzheimer's disease. | PET study only. |
| Ismail et al. | A functional neuroimaging study of appetite loss in Alzheimer's disease. | SPECT study only. |
| Lanctot et al. | A SPECT study of apathy in alzheimer's disease | SPECT study only. |
| Callen et al. | Limbic system perfusion in Alzheimer's disease measured by MRI-coregistered HMPAO SPET | SPECT study only. |
| Lanctot et al. | Medial Temporal Hypoperfusion and Aggression in Alzheimer Disease. | SPECT study only. |
